# Asymptomatic SARS-CoV-2 infections: a living systematic review and meta-analysis

**DOI:** 10.1101/2020.04.25.20079103

**Authors:** Diana Buitrago-Garcia, Dianne Egli-Gany, Michel J Counotte, Stefanie Hossmann, Hira Imeri, Mert Aziz Ipekci, Georgia Salanti, Nicola Low

**Affiliations:** Institute of Social and Preventive Medicine, University of Bern, Bern, Switzerland; Clinical Trials Unit, University of Bern, Bern, Switzerland

## Abstract

**BACKGROUND:** There is disagreement about the level of asymptomatic severe acute respiratory syndrome coronavirus 2 (SARS-CoV-2) infection. We conducted a living systematic review and meta-analysis to address three questions: 1. amongst people who become infected with SARS-CoV-2, what proportion does not experience symptoms at all during their infection? 2. Amongst people with SARS-CoV-2 infection who are asymptomatic when diagnosed, what proportion will develop symptoms later? 3. What proportion of SARS-CoV-2 transmission is accounted for by people who are either asymptomatic throughout infection, or pre-symptomatic?

**METHODS AND FINDINGS:** We searched PubMed, Embase, bioRxiv and medRxiv using a database of SARS-CoV-2 literature that is updated daily, on 25 March 2020, 20 April 2020 and 10 June 2020. Studies of people with SARS-CoV-2 diagnosed by reverse transcriptase PCR that documented follow-up and symptom status at the beginning and end of follow-up, or modelling studies were included. One reviewer extracted data and a second verified the extraction, with disagreement resolved by discussion or a third reviewer. Risk of bias in empirical studies was assessed with an adapted checklist for case series and the relevance and credibility of modelling studies were assessed using a published checklist. We included a total of 94 studies. The overall estimate of the proportion of people who become infected with SARS-CoV-2 and remain asymptomatic throughout infection was 20% (95% CI 17-25) with a prediction interval of 3-67% in 79 studies that addressed this review question. There was some evidence that biases in the selection of participants influence the estimate. In seven studies of defined populations screened for SARS-CoV-2 and then followed, 31% (95% CI 26-37%, prediction interval 24-38%) remained asymptomatic. The proportion of people that is pre-symptomatic could not be summarised, owing to heterogeneity. The secondary attack rate was slightly lower in contacts of people with asymptomatic infection than those with symptomatic infection (relative risk 0.35, 95% CI 0.10-1.27). Modelling studies fit to data found a higher proportion of all SARS-CoV-2 infections resulting from transmission from pre-symptomatic individuals than from asymptomatic individuals. Limitations of the review include that most included studies were not designed to estimate the proportion of asymptomatic SARS-CoV-2 infections and were at risk of selection biases, we did not consider the possible impact of false negative RT-PCR results, which would underestimate the proportion of asymptomatic infections, and that the database does not include all sources.

**CONCLUSIONS:** The findings of this living systematic review of publications early in the pandemic suggest that most SARS-CoV-2 infections are not asymptomatic throughout the course of infection. The contribution of pre-symptomatic and asymptomatic infections to overall SARS-CoV-2 transmission means that combination prevention measures, with enhanced hand hygiene, masks, testing tracing and isolation strategies and social distancing, will continue to be needed.

**AUTHOR SUMMARY:** *Why was this study done?:*

▪ The proportion of people who will remain asymptomatic throughout the course of infection with severe acute respiratory syndrome coronavirus 2 (SARS-CoV-2), the cause of coronavirus disease 2019 (covid-19), is not known.
▪ Studies that assess people at just one time point will overestimate the proportion of true asymptomatic infection because those who go on to develop covid-19 symptoms will be wrongly classified as asymptomatic, rather than pre-symptomatic.
▪ The amount, and infectiousness, of asymptomatic SARS-CoV-2 infection will determine what kind of measures will prevent transmission most effectively. What did the researchers do and find?

▪ We did a living systematic review through 10 June 2020, using automated workflows that speed up the review processes, and allow the review to be updated when relevant new evidence becomes available.
▪ Overall, in 79 studies in a range of different settings, 20% (95% confidence interval, CI 17–25%) of people with SARS-CoV-2 infection remained asymptomatic during follow-up, but biases in study designs limit the certainty of this estimate.
▪ We found some evidence that SARS-CoV-2 infection in contacts of people with asymptomatic infection is less likely than in contacts of people with symptomatic infection (relative risk 0.35, 95% CI 0.10-1.27). What do these findings mean?

▪ The findings of this living systematic review suggest that most SARS-CoV-2 infections are not asymptomatic throughout the course of infection.
▪ Future studies should be designed specifically to determine the true proportion of asymptomatic SARS-CoV-2 infections, using methods to minimise biases in the selection of study participants and ascertainment of symptom status during follow up.
▪ The contribution of pre-symptomatic and asymptomatic infections to overall SARS-CoV-2 transmission means that combination prevention measures, with enhanced hand hygiene, masks, testing tracing and isolation strategies and social distancing, will continue to be needed.

**Changes from version 2:**

▪ Search updated 10.06.2020, total number of included studies increased from 37 to 94
▪ Protocol updated at https://osf.io/9ewys/
▪ New analyses
  - Review question 1, prediction intervals added for each study setting
  - Meta-analysis of secondary attack rate from asymptomatic and pre-symptomatic index cases compared with symptomatic
  - Sensitivity analysis for review question 1, omitting preprints
▪ Conclusions unchanged

## Introduction

There is ongoing discussion about the level of asymptomatic severe acute respiratory syndrome coronavirus 2 (SARS-CoV-2) infection. The authors of a narrative review report a range of proportions of participants positive for SARS-CoV-2 but asymptomatic in different studies from 6 to 96% [1]. The discrepancy results, in part, from the interpretation of studies that report a proportion of asymptomatic people with SARS-CoV-2 detected at a single point. The studies cited include both people who will remain asymptomatic throughout and those, known as pre-symptomatic, who will develop symptoms of coronavirus disease 2019 (covid-19) if followed up [2]. The full spectrum and distribution of covid-19, from completely asymptomatic, to mild and non-specific symptoms, viral pneumonia, respiratory distress syndrome, and death are not yet known [3]. Without follow up, however, the proportions of asymptomatic and pre-symptomatic infections cannot be determined.

Accurate estimates of the proportions of true asymptomatic and pre-symptomatic infections are needed urgently because their contribution to overall SARS-CoV-2 transmission at the population level will determine the appropriate balance of control measures [3]. If the predominant route of transmission is from people who have symptoms, then strategies should focus on testing, followed by isolation of infected individuals and quarantine of their contacts. If, however, most transmission is from people without symptoms, social distancing measures that reduce contact with people who might be infectious, should be prioritised, enhanced by active case-finding through testing of asymptomatic people.

The objectives of this study were to address three questions: 1. Amongst people who become infected with SARS-CoV-2, what proportion does not experience symptoms at all during their infection? 2. Amongst people with SARS-CoV-2 infection who are asymptomatic when diagnosed, what proportion will develop symptoms later? 3. What proportion of SARS-CoV-2 transmission is accounted for by people who are either asymptomatic throughout infection, or pre-symptomatic?

## Methods

We conducted a living systematic review, a systematic review that provides an online summary of findings and is updated when relevant new evidence becomes available [4]. The review follows a published protocol (https://osf.io/9ewys/), which describes in detail the methods used to speed up review tasks [5] and to assess relevant evidence rapidly during a public health emergency [6]. The first two versions of the review have been published as preprints [7,8]. We report our findings according to the statement on preferred reporting items for systematic reviews and meta-analyses (S1 PRISMA Checklist) [9]. Box 1 shows our definitions of symptoms, asymptomatic infection and pre-symptomatic status. We use the term asymptomatic SARS-CoV-2 infection for people without symptoms of covid-19 who remain asymptomatic throughout the course of infection. We use the term pre-symptomatic for people who do not have symptoms of covid-19 when enrolled in a study, but who develop symptoms during adequate follow-up.

**Box 1**

**Definitions of symptoms and symptom status in a person with SARS-CoV-2 infections**

#### Symptoms

symptoms that a person experiences and reports. We used the authors’ definitions. We searched included manuscripts for an explicit statement that the study participant did not report symptoms that they experienced. Some authors defined ‘asymptomatic’ as an absence of self-reported symptoms. We did not include clinical signs observed or elicited on examination.

#### Asymptomatic infection

a person with laboratory-confirmed SARS-CoV-2 infection, who has no symptoms, according to the authors’ report, at the time of first clinical assessment and had no symptoms at the end of follow-up. The end of follow-up was defined as any of the following: virological cure, with one or more negative RT-PCR test results; follow-up for 14 days or more after the last possible exposure to an index case; follow-up for seven days or more after the first RT-PCR positive result.

#### Pre-symptomatic

a person with laboratory-confirmed SARS-CoV-2 infection, who has no symptoms, according to the authors’ report, at the time of first clinical assessment, but who developed symptoms by the end of follow-up. The end of follow-up was defined as any of the following: virological cure, with one or more negative RT-PCR test results; follow-up for 14 days or more after the last possible exposure to an index case; follow-up for seven days or more after the first RT-PCR positive result.

### Information sources and search

We conducted the first search on March 25, 2020 and updated it on April 20 and June 10, 2020. We searched the covid-19 living evidence database [10], which is generated using automated workflow processes [5] to: i) provide daily updates of searches of four electronic databases: Medline Pubmed, Ovid Embase, bioRxiv and medRxiv, using medical subject headings and free text keywords for SARS-CoV-2 infection and covid-19; ii) de-duplicate the records; iii) tag records that are preprints; and iv) allow searches of titles and abstracts using Boolean operators. We used the search function to identify studies of asymptomatic or pre-symptomatic SARS-CoV-2 infection using a search string of medical subject headings and free text keywords (supporting information, S1 Text). We also examined articles suggested by experts and the reference lists of retrieved mathematical modelling studies and systematic reviews. Reports from this living rapid systematic review will be updated at three-monthly intervals, with continuously updated searches.

### Eligibility criteria

We included studies in any language of people with SARS-CoV-2 diagnosed by reverse transcriptase PCR (RT-PCR) that documented follow-up and symptom status at the beginning and end of follow-up, or investigated the contribution to SARS-CoV-2 transmission of asymptomatic or pre-symptomatic infection. We included contact tracing investigations, case series, cohort studies, case-control studies and statistical and mathematical modelling studies. We excluded the following study types: case reports of a single patient and case series where participants were not enrolled consecutively. Where multiple records included data from the same study population, we linked the records and extracted data from the most complete report.

### Study selection and data extraction

Reviewers worked in pairs to screen records using an application programming interface in the electronic data capture system (REDCap, Vanderbilt University, USA). One reviewer selected potentially eligible studies and a second reviewer verified all included and excluded studies. We reported the identification, exclusion and inclusion of studies in a flowchart (S1 Figure). The reviewers determined which of the three review questions each study addressed, using the definitions in Box 1. One reviewer extracted data using a pre-piloted extraction form in REDCap and a second reviewer verified the extracted data using the query system. A third reviewer adjudicated on disagreements that could not be resolved by discussion. We contacted study authors for clarification where the study description was insufficient to reach a decision on inclusion or if reported data in the manuscript were internally inconsistent. The extracted variables included, but were not limited to, study design, country and/or region, study setting, population, age, primary outcomes and length of follow-up. From empirical studies, we extracted raw numbers of individuals with any outcome and its relevant denominator. From statistical and mathematical modelling studies we extracted proportions and uncertainty intervals reported by the authors.

The primary outcomes for each review question were: 1. Proportion with asymptomatic SARS-CoV-2 infection who did not experience symptoms at all during follow-up; 2. Proportion with SARS-CoV-2 infections who did not have symptoms at the time of testing but developed symptoms during follow-up. 3. Estimated proportion (with uncertainty interval) of SARS-CoV-2 transmission accounted for by people who are asymptomatic or pre-symptomatic. A secondary outcome for review question 3 was the secondary attack rate from asymptomatic or pre-symptomatic index cases.

### Risk of bias in included studies

Two authors independently assessed the risk of bias. A third reviewer resolved disagreements. For observational epidemiological studies, we adapted the Joanna Briggs Institute Critical Appraisal Checklist for Case Series [11]. The adapted tool included items about inclusion criteria, measurement of asymptomatic status, follow-up of course of disease, and statistical analysis. We added items about selection biases affecting the study population from a tool for the assessment of risk of bias in prevalence studies [12]. For mathematical modelling studies, we used a checklist for assessing relevance and credibility [13].

### Synthesis of the evidence

We used the *metaprop* and *metabin* functions from the *meta* package (version 4.11-0) [14] in R (version 3.5.1) to display the study findings in forest plots and synthesise their findings. The 95% confidence intervals (CI) for each study are estimated using the Clopper-Pearson method [15]. We examined heterogeneity visually in forest plots. We stratified studies according to the methods used to identify people with asymptomatic SARS-CoV-2 infection and the study setting. To synthesise proportions from comparable studies, in terms of design and population, we used stratified random effects meta-analysis. For the stratified and overall summary estimates we calculated prediction intervals, to represent the likely range of proportions that would be obtained in subsequent studies conducted in similar settings [16]. We calculated the secondary attack rate as the number of cases among contacts as a proportion of all close contacts ascertained. We did not account for potential clustering of contacts because the included studies did not report the size of clusters. We compared the secondary attack rate from asymptomatic or pre-symptomatic index cases with that from symptomatic cases. If there were no events in a group, we added 0.5 to each cell in the 2×2 table. We used random effects meta-analysis with the Mantel-Haenszel method to estimate a summary risk ratio (with 95% CI).

## Results

The living evidence database contained a total of 25538 records about SARS-CoV-2 or COVID-19 by 10 June, 2020. The searches for studies about asymptomatic or pre-symptomatic SARS-CoV-2, on 25 March, 20 April and 10 June, resulted in 89, 230 and 688 records for screening (S1 Figure). In the first version of the review [7], 11 articles were eligible for inclusion [17-27], version 2 [8] identified another 26 eligible records [28-53], and version 3 identified another 61 eligible records [54-114]. After excluding four articles for which more recent data became available in a subsequent version [25,29,30,35], the total number of articles included was 94 (S1 Table) [17-24,26-28,31-34,36-114]. The types of evidence changed across the three versions of the review (S1 Table). In the first version, six of 11 studies were contact investigations of single family clusters with a total of 39 people. In the next versions, study designs included larger investigations of contacts and outbreaks, screening of defined groups and studies of hospitalised adults and children. Across all three review versions, data from 79 empirical observational studies were collected in 19 countries or territories (Tables 1 and 2) and included 6832 people with SARS-CoV-2 infection. Forty seven of the studies, including 3802 infected people were done in China (S2 Table). At the time of their inclusion in the review, 23 of the included records were preprints; six of these had been published in peer-reviewed journals by 17 July 2020 [19,20,27,81,82,106].

**Table 1.**
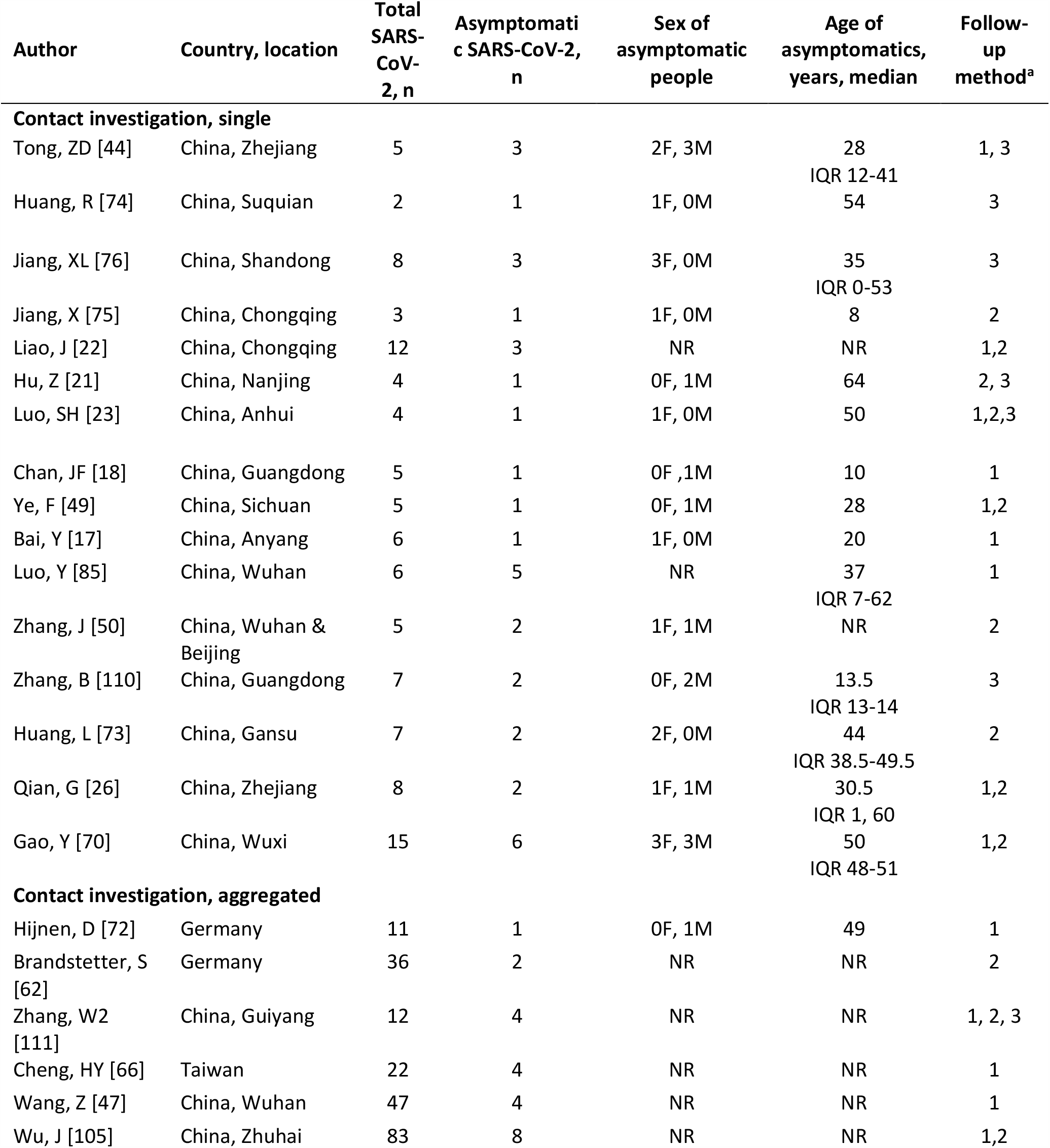
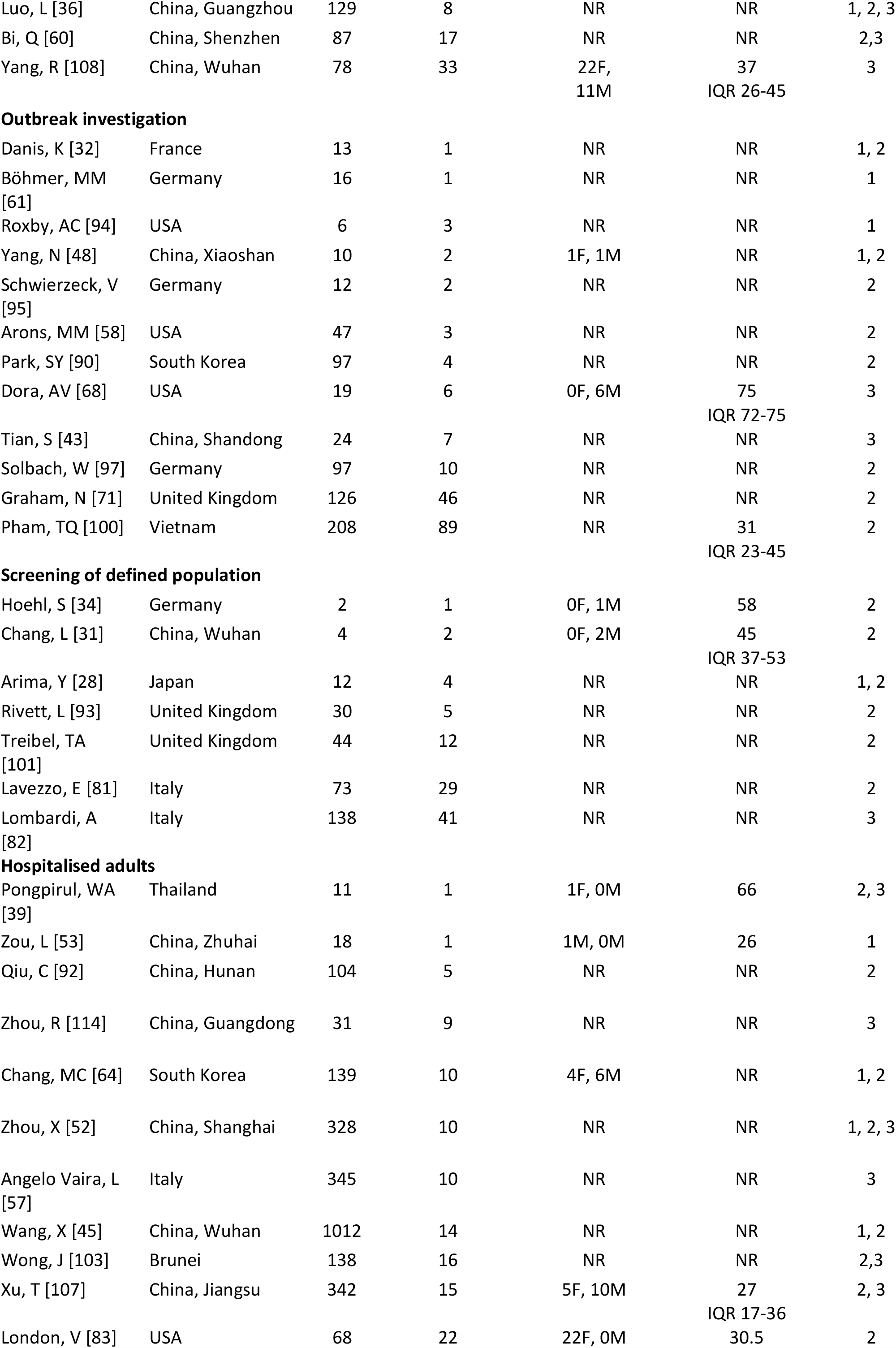
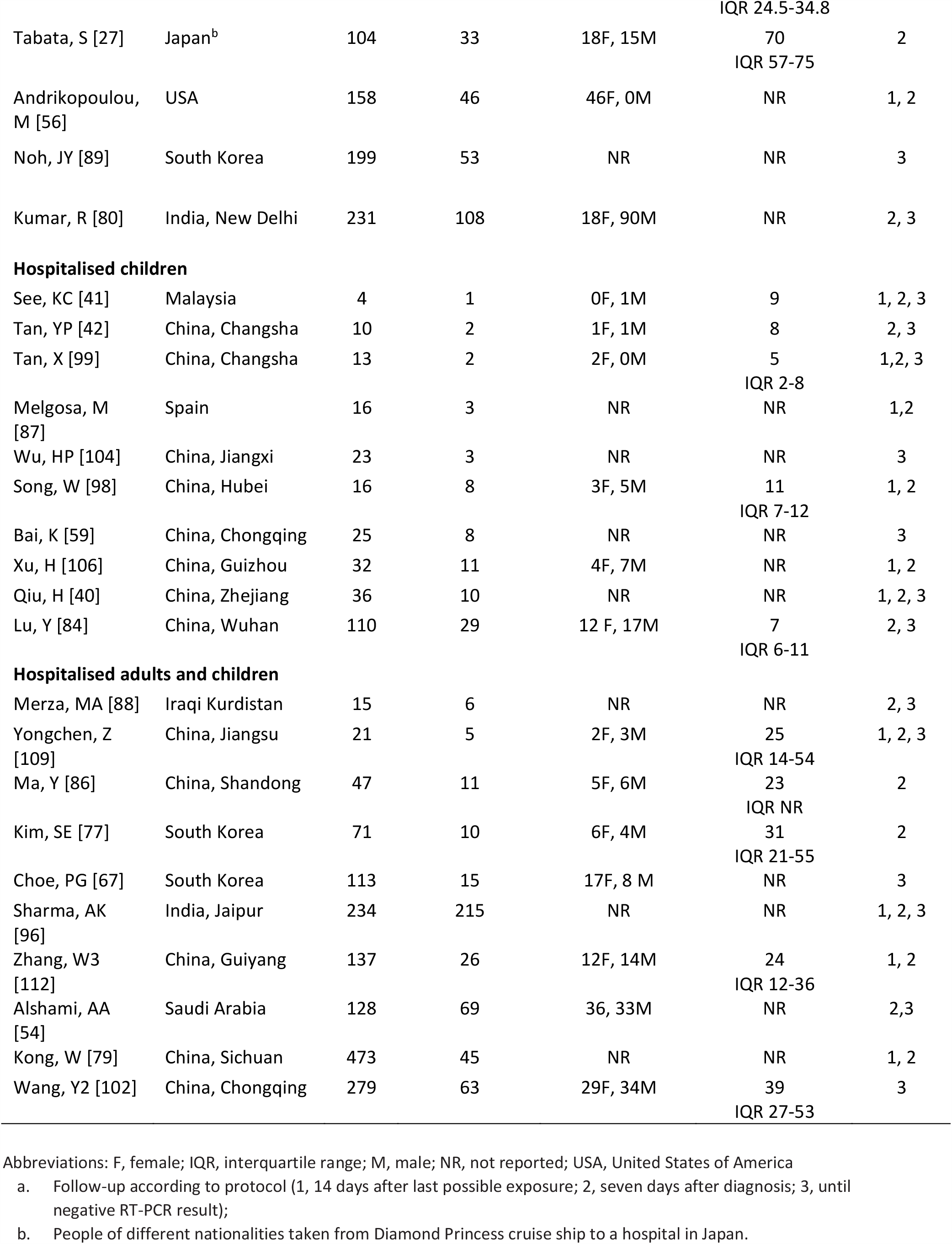
Characteristics of studies reporting on proportion of asymptomatic SARS-CoV-2 infections

**Table 2.** Characteristics of studies that measured the proportion of people with SARS-CoV-2 infection that develops symptoms

| Author | Country, location | Total asymptomatic SARS-CoV-2, n | Develops symptoms after testing, n | Sex, asymptomatics at time of testing | Age of asymptomatics at time of testing, years, median | Follow-up method<sup>a</sup> |
| --- | --- | --- | --- | --- | --- | --- |
| **Contact investigation, single** |  |  |  |  |  |  |
| Ye, F [49] | China, Sichuan | 3 | 2 | 0F, 3M | 28<br>IQR 23-50 | 1, 2 |
| Zhang, B [110] | China, Guangdong | 4 | 2 | 0F, 4M | 34<br>IQR 33-35 | 3 |
| Huang, L [73] | China, Gansu | 4 | 2 | 3F, 1M | 44.5<br>IQR 34.50-54.25 | 2 |
| Jiang, XL [76] | China, Shandong | 5 | 2 | 3F, 2M | 35<br>IQR 35-37 | 3 |
| Hu, Z [21] | China, Nanjing | 24 | 5 | NR | NR | 2, 3 |
| **Contact investigation, aggregated** |  |  |  |  |  |  |
| Zhang, W2 [111] | China, Guangzhou | 12 | 8 | NR | NR | 1,2,3 |
| **Outbreak investigation** |  |  |  |  |  |  |
| Schwiezeck, V [95] | Germany | 6 | 4 | NR | NR | 2 |
| Park, SY [90] | South Korea | 8 | 4 | NR | NR | 2 |
| Arons, MM [58] | USA | 27 | 24 | NR | NR | 2 |
| Dora, AV [68] | USA | 14 | 8 | 0F, 14M | NR | 3 |
| Graham, N [71] | United Kingdom | 54 | 8 | NR | NR | 1 |
| **Screening of defined population** |  |  |  |  |  |  |
| Hoehl, S [34] | Germany | 2 | 1 | 1F, 1M | 51 | 2 |
| Rivett, L [93] | United Kingdom | 6 | 1 | NR | NR | 2 |
| Chang, L [31] | China, Wuhan | 4 | 2 | 1F, 3M | 39.5<br>IQR 29-47.5 | 2 |
| Arima, Y [28] | Japan | 5 | 2 | NR | NR | 1, 2 |
| Lytras, T [37] | Greece | 39 | 4 | NR | NR | 2 |
| Lavezzo, E [81] | Italy | 39 | 10 | NR | NR | 2 |
| **Hospitalised adults** |  |  |  |  |  |  |
| Al-Shamsi, HO [55] | United Arab Emirates | 7 | 7 | 5F, 2M | 51.6<br>IQR 40-76 | 3 |
| Luo, SH [23] | China, Anhui | 8 | 7 | NR | NR | 1, 2, 3 |
| Zhou, X [52] | China, Shanghai | 13 | 3 | 7F, 6M | NR | 2, 3 |
| Zhou, R [114] | China, Guangdong | 31 | 22 | NR | NR | 3 |
| Wang, X [45] | China, Wuhan | 30 | 16 | NR | NR | 1, 2 |
| Tabata, S [27] | Cruise Ship | 43 | 10 | 24F, 19M | 69<br>IQR 60.5-75 | 2 |
| Wang, Y1 [46] | China, Shenzhen | 55 | 43 | NR | 49<br>IQR 2-69 | 3 |
| Meng, H [38] | China, Wuhan | 58 | 16 | NR | NR | 2 |
| Andrikopoulou, M [56] | USA | 63 | 16 | 63F, 0M | NR | 1, 2 |
| Zhang, Z [113] | China, Shenzhen | 56 | 33 | 33F, 23M | NR | 2,3 |
| Wong, J [103] | Brunei | 138 | 42 | NR | NR | 2, 3 |
| **Hospitalised children** |  |  |  |  |  |  |
| See, KC [41] | Malaysia | 2 | 1 | 0F, 2M | 5<br>IQR 1-9 | 1, 2, 3 |
| **Hospitalised adults and children** |  |  |  |  |  |  |
| Kim, SE [77] | South Korea | 13 | 3 | 7F, 6M | 31<br>IQR 20.5-51.5 | 2 |
| Choe, PG [67] | South Korea | 54 | 39 | 32F, 22M | NR | 3 |
| Kong, W [79] | China, Sichuan | 62 | 17 | NR | NR | 1 |

Abbreviations: F, female; IQR, interquartile range; M, male; NR, not reported; USA, United States of America
- a. Follow-up according to protocol (1, 14 days after possible exposure; 2, seven days after diagnosis; 3, until one or more negative RT-PCR result); - b. People of different nationalities taken from Diamond Princess cruise ship to a hospital in Japan - c. Until hospital discharge or negative RT-PCR.

### Proportion of people with asymptomatic SARS-CoV-2 infection

We included 79 studies that reported empirical data about 6616 people with SARS-CoV-2 infection (1287 defined as having asymptomatic infection) [17,18,21-23,26-28,31,32,34,36,39-45,47-50,52-54,56-62,64,66-68,70-77,79-90,92-112,114] and one statistical modelling study [24] (Table 1). The sex distribution of the people with asymptomatic infection was reported in 41/79 studies and the median age was reported in 35/79 studies (Table 1). The results of the studies were heterogeneous (S2 Figure). We defined seven strata, according to the method of selection of asymptomatic status and study settings. Study findings within some of these strata were more consistent (Figure 1). We considered the statistical modelling study of passengers on the Diamond Princess cruise ship passengers [24] separately, because of the different method of analysis and overlap with the study population reported by Tabata S, et al. [27].

**Figure 1.**
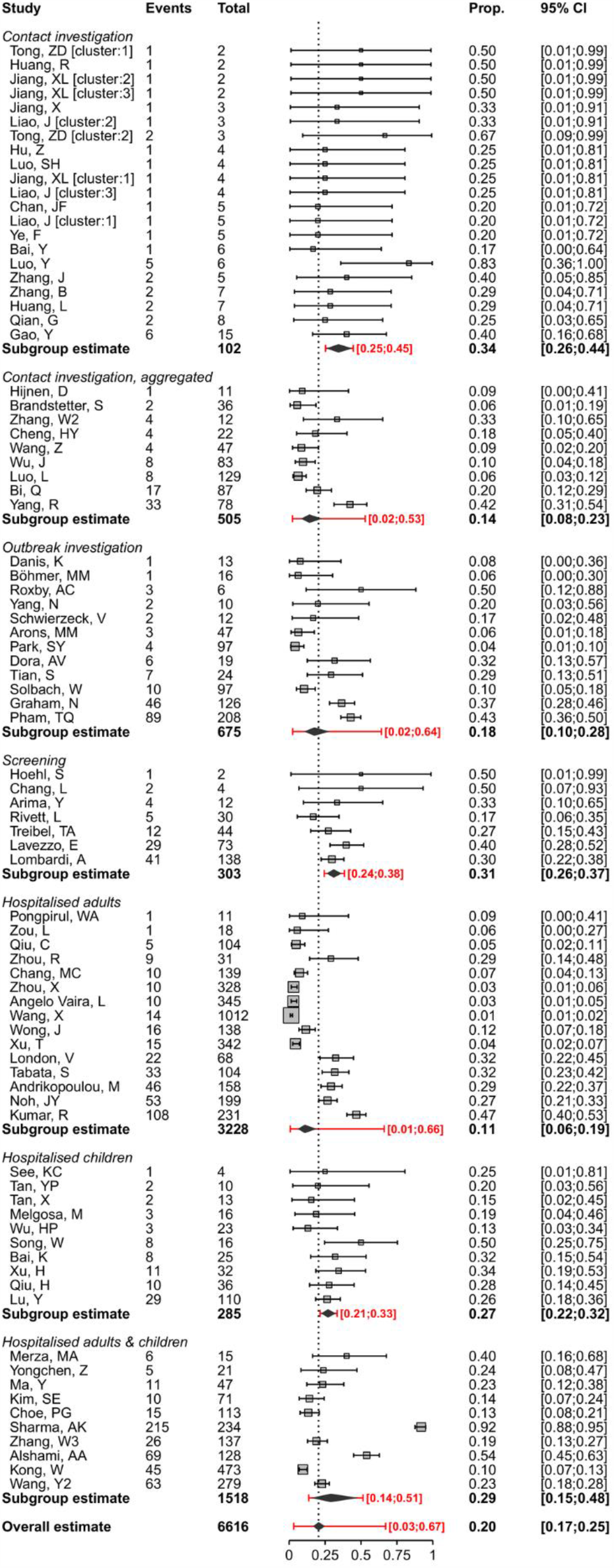
Forest plot of proportion of people with asymptomatic SARS-CoV-2 infection, stratified by setting. The x-axis displays proportions. In the setting ‘Contact investigations’ where more than one cluster was reported, clusters are annotated with ‘[cluster]’. The diamond shows the summary estimate and its 95% confidence interval. The red bar and red text show the prediction interval.

The main risks of bias across all categories of empirical studies were in the selection and enrolment of people with asymptomatic infection and mismeasurement of asymptomatic status because of absent or incomplete definitions (S3 Figure). Sources of bias specific to studies in particular settings are discussed with the relevant results.

The overall estimate of the proportion of people who become infected with SARS-CoV-2 and remain asymptomatic throughout the course of infection was 20% (95% CI 17–25%, 79 studies), with a prediction interval of 3 to 67% (Figure 1). One statistical modelling study was based on data from all 634 passengers from the Diamond Princess Cruise ship with RT-PCR positive test results [24]. The authors adjusted for the proportion of people who would develop symptoms (right censoring) in a Bayesian framework to estimate that, if all were followed up until the end of the incubation period, the probability of asymptomatic infections would be 17.9% (95% credibility interval, CrI 15.5–20.2%).

The summary estimates of the proportion of people with asymptomatic SARS-CoV-2 infection differed according to study setting, although prediction intervals for all groups overlapped. The first three strata in Figure 1 involve studies that reported on different types of contact investigation, which start with an identified covid-19 case. The studies reporting on single family clusters (21 estimates from 16 studies in China, n=102 people with SARS-CoV-2) all included at least one asymptomatic person [17,18,21-23,26,44,49,50,70,73-76,85,110]. The summary estimate was 34% (95% CI 26–44%, prediction interval 25–45%). In nine studies that reported on close contacts of infected individuals and aggregated data from clusters of both asymptomatic and symptomatic people with SARS-CoV-2 the summary estimate was 14% (95% CI 8–23%, prediction interval 2–53%) [36,47,60,62,66,72,105,108,111]. We included 12 studies (n=675 people) that reported on outbreak investigations arising from a single symptomatic person, or from the country’s first imported cases of people with covid-19 [32,43,48,58,61,68,71,90,94,95,97,100]. Four of the outbreaks involved nursing homes [58,68,71,94] and four involved occupational settings [43,61,90,95]. The summary estimate of the proportion of asymptomatic SARS-CoV-2 infections was 18% (95% CI 10–28%, prediction interval 10–28%).

In seven studies, people with SARS-CoV-2 infection were detected through screening of all people in defined populations who were potentially exposed (303 infected people amongst 10090 screened) [28,31,34,81,82,93,101]. The screened populations included healthcare workers [82,93,101], people evacuated from a setting where SARS-CoV-2 transmission was confirmed, irrespective of symptom status [28,34], the whole population of one village in Italy [81] and blood donors [31]. In these studies, the summary estimate of the proportion asymptomatic was 31% (95% CI 26–37%, prediction interval 24–38%). There is a risk of selection bias in studies of certain groups, such as healthcare workers and blood donors, because people with symptoms are excluded [31,82,93,101] or from non-responders in population-based screening [81]. Retrospective symptom ascertainment could also increase the proportion determined asymptomatic [81,82,101].

The remaining studies, in hospital settings, included adult patients only (15 studies, n=3228) [27,39,45,52,53,56,57,64,80,83,89,92,103,107,114], children only (10 studies, n=285) [40-42,59,84,87,98,99,104,106] or adults and children (10 studies, n=1518) [54,67,77,79,86,88,96,102,109,112] (Table 1, Figure 1). The types of hospital and clinical severity of patients differed, including settings in which anyone with SARS-CoV-2 infection was admitted for isolation and traditional hospitals.

### Proportion of pre-symptomatic SARS-CoV-2 infections

We included 31 studies in which the people with no symptoms of covid-19 at enrolment were followed up and the proportion that develops symptoms is defined as pre-symptomatic (Table 2, Figure 2) [21,27,28,31,34,37,38,41,45,46,49,52,55,56,58,67,68,71,73,76,77,79,81,90,93,95,103,110,111,113,114]. Four studies addressed only this review question [37,38,55,113]. The findings from the 31 studies were heterogeneous (S4 Figure), even when categorised according to the method of selection of asymptomatic participants, and we did not estimate a summary measure (Figure 2).

**Figure 2.**
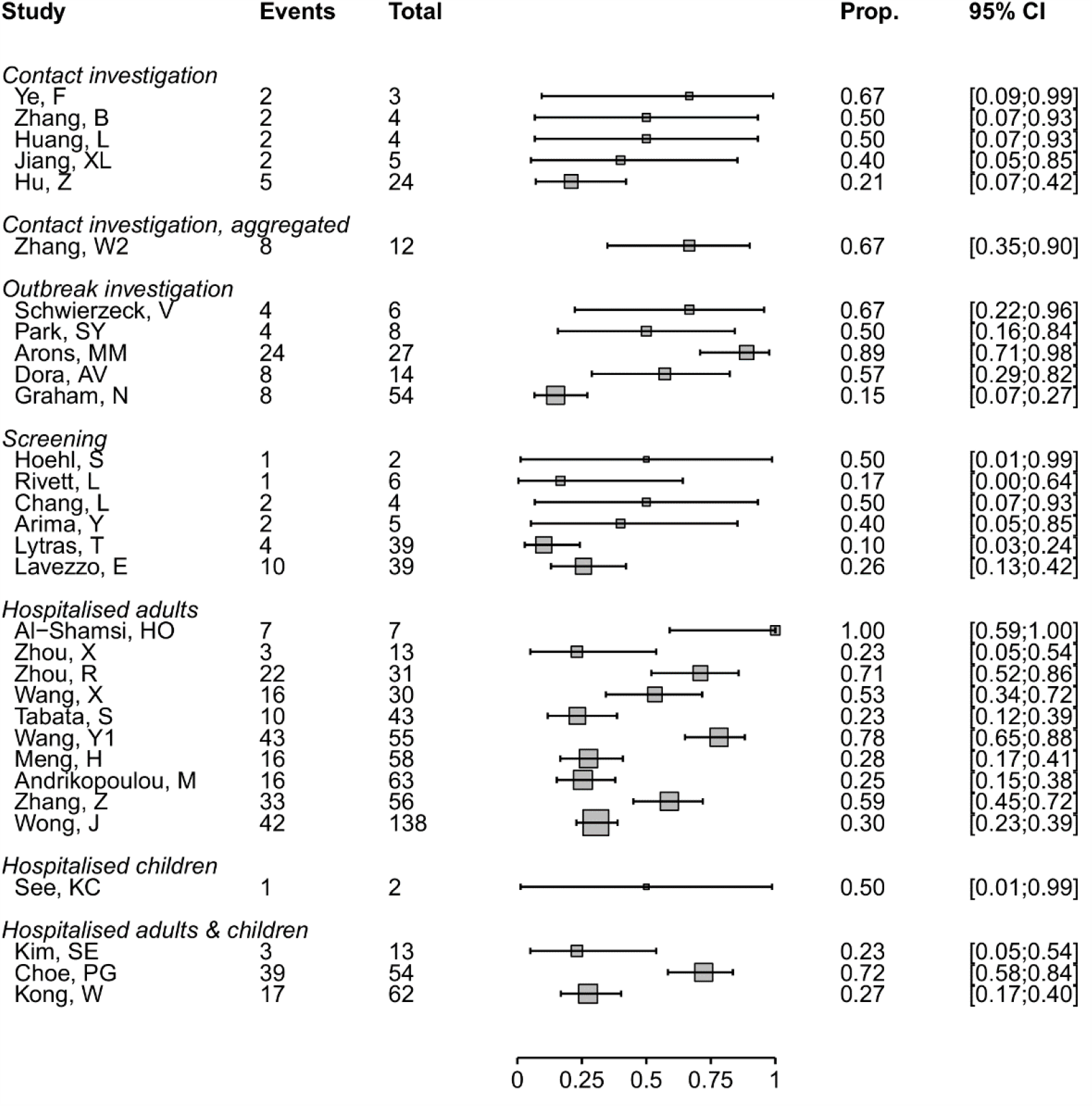
Forest plot of proportion of people with pre-symptomatic SARS-CoV-2 infection, stratified by setting. The x-axis displays proportions.

### Additional analyses

We investigated heterogeneity in the estimates of the proportion of asymptomatic SARS-CoV-2 infections in subgroup analyses that were not specified in the original protocol. In studies of hospitalised children, the point estimate was higher (25%, 95% CI 14–40%, 10 studies) than in adults (11%, 95% CI 7–17%, 15 studies), but confidence intervals overlapped (Figure 1). The proportion of asymptomatic SARS-CoV-2 infection estimated in studies of hospitalised patients (35 studies, 19%, 95% CI 14–25%) was similar to that in all other settings (44 studies, 22%, 95% CI 17–29%, S5 Figure). To examine publication status, we conducted a sensitivity analysis, omitting studies that were identified as preprints at the time of data extraction (S6 Figure). The estimate of the proportion of asymptomatic infection in all settings (18%, 95% CI 14–22%) and setting-specific estimates were very similar to the main analysis.

### Contribution of asymptomatic and pre-symptomatic infection to SARS-CoV-2 to transmission

Five of the studies that conducted detailed contact investigations provided enough data to calculate a secondary attack rate according to the symptom status of the index cases (Figure 3) [36,65,66,90,111]. The summary risk ratio for asymptomatic compared with symptomatic was 0.35 (95% CI 0.1–1.27) and for pre-symptomatic compared with symptomatic people was 0.63 (95% CI 0.18–2.26) [66,90]. The risk of bias in ascertainment of contacts was judged to be low in all studies.

**Figure 3.**
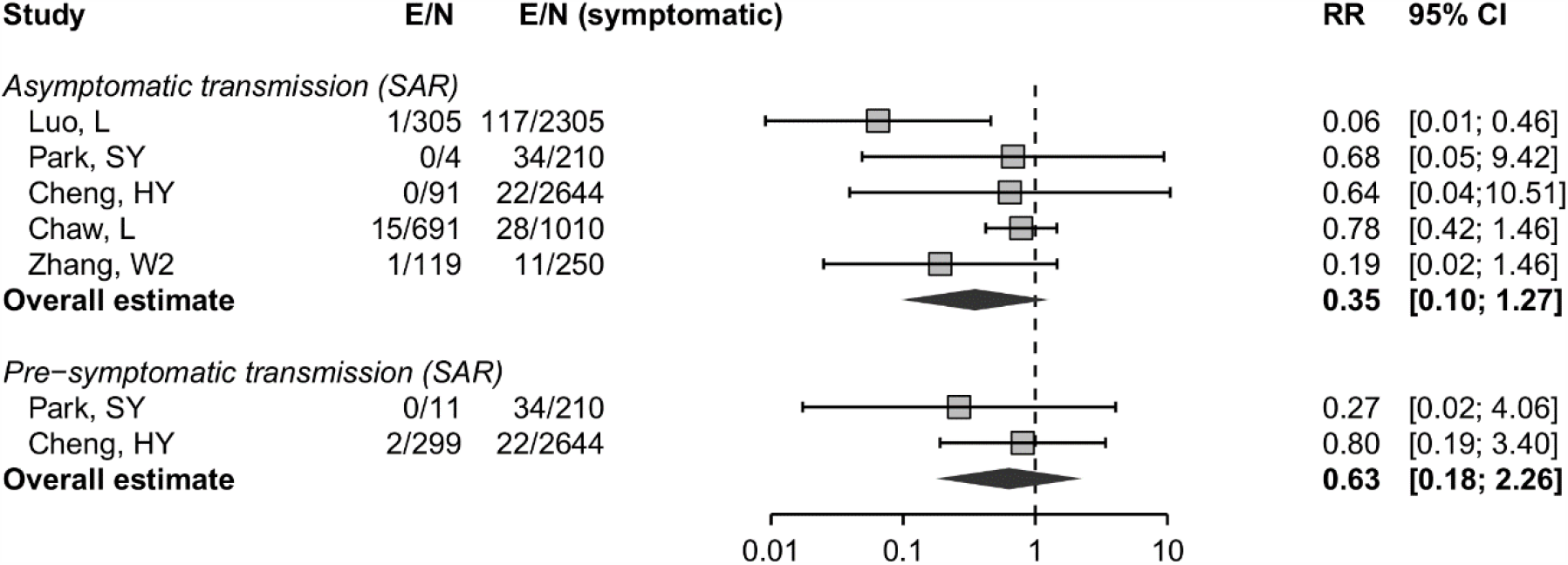
Forest plot of the risk ratio (RR) and 95% confidence interval (CI) of the secondary attack rate (SAR), comparing infections in contacts of asymptomatic and pre-symptomatic index cases with infections in contacts of symptomatic cases. E, number of secondary transmission events; N, number of close contacts. The x-axis shows the risk ratio on a logarithmic scale.

We included eight mathematical modelling studies (Figure 4) [19,20,33,51,63,69,78,91]. The models in five studies were informed by analysis of data from contact investigations in China, South Korea, Singapore, and the Diamond Princess cruise ship, using data to estimate the serial interval or generation time [19,20,33,69,78] and in three studies the authors used previously published estimates [51,63,91].

**Figure 4.**
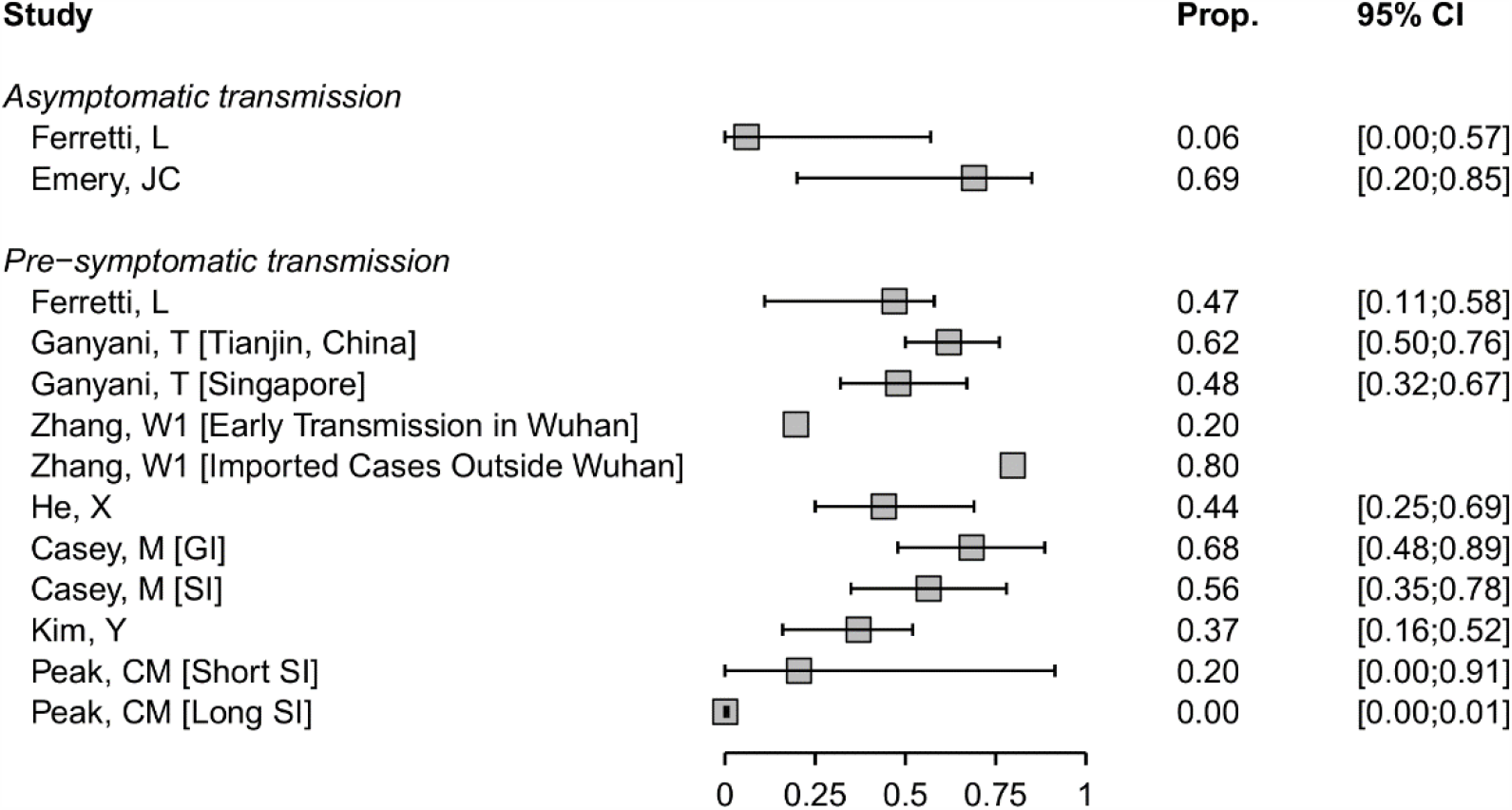
Forest plot of proportion of SARS-CoV-2 infection resulting from asymptomatic or pre-symptomatic transmission. For studies that report outcomes in multiple settings, these are annotated in brackets. SI, serial interval; GI: generation interval.

Estimates of the contributions of both asymptomatic and pre-symptomatic infections SARS-CoV-2 transmission were very heterogeneous. In two studies, the contribution to SARS-CoV-2 transmission of asymptomatic infection were estimated to be 6% (95% CrI 0–57%) [19] and 69% (95% CrI 20–85%) [69] (Figure 4). The estimates have large uncertainty intervals and the disparate predictions result from differences in the proportion of asymptomatic infections and relative infectiousness of asymptomatic infection. Ferretti L, et al. provide an interactive web application [ref:[link](https://bdi-pathogens.shinyapps.io/covid-19-transmission-routes/)], which shows how these parameters affect the model results.

Models of the contribution of pre-symptomatic transmission used different assumptions about the durations and distributions of infection parameters such as incubation period, generation time and serial interval [19,20,33,51,63,78,91]. In models that accounted for uncertainty appropriately, most estimates of the proportion of transmission resulting from people with SARS-CoV-2 who are pre-symptomatic ranged from 20 to 70%. In one study that estimated a contribution of <1% [91], the model fitted serial interval was longer than observed in empirical studies [115]. The credibility of most modelling studies was limited by the absence of external validation. The data to which the models were fitted were generally from small samples (S7 Figure).

## Discussion

### Summary of main findings

The summary proportion of SARS-CoV-2 that is asymptomatic throughout the course of infection was estimated to be 20% (95% CI 17–25%, 79 studies), with a prediction interval of 3–67%. In studies that identified SARS-CoV-2 infection through screening of defined populations, the proportion of asymptomatic infections was 31% (95% CI 26–37%, 7 studies). In 31 studies reporting on people who are pre-symptomatic but who go on to develop symptoms, the results were too heterogeneous to combine. The secondary attack rate from asymptomatic infections may be lower than that from symptomatic infections (relative risk 0.35, 95% CI 0.1–1.27). Modelling studies estimated a wide range of the proportion of all SARS-CoV-2 infections that result from transmission from asymptomatic and pre-symptomatic individuals.

### Strengths and weaknesses

A strength of this review is that we used clear definitions and separated review questions to distinguish between SARS-CoV-2 infections that remain asymptomatic throughout their course from those that become symptomatic, and to separate proportions of people with infection from their contribution to transmission in a population. This living systematic review uses methods to minimise bias whilst increasing the speed of the review process [5,6], and will be updated regularly. We only included studies that provided information about follow-up through the course of infection, which allowed reliable assessment about the proportion of asymptomatic people in different settings. In the statistical synthesis of proportions, we used a method that accounts for the binary nature of the data and avoids the normality approximation (weighted logistic regression).

Limitation of the review are that most included studies were not designed to estimate the proportion of asymptomatic SARS-CoV-2 infection and definitions of asymptomatic status were often incomplete or absent. The risks of bias, particularly those affecting selection of participants, differed between studies and could result in both underestimation and overestimation of the true proportion of asymptomatic infections. Also, we did not consider the possible impact of false negative RT-PCR results, which might be more likely to occur in asymptomatic infections [116] and would underestimate the proportion of asymptomatic infections [117]. The four databases that we searched are not comprehensive, but they cover the majority of publications and we do not believe that we have missed studies that would change our conclusions.

### Comparison with other reviews

We found narrative reviews that reported wide ranges (five to 96%) of infections that might be asymptomatic [1,118]. These reviews presented cross-sectional studies alongside longitudinal studies and did not distinguish between asymptomatic and pre-symptomatic infection. We found three systematic reviews, which reported similar summary estimates from meta-analysis of studies published up to May [119-121]. In two reviews, authors applied inclusion criteria to reduce the risks of selection bias, with summary estimates of 11% (95% CI 4–18%, 6 studies) [120] and 15% (95% CI 12–18%, 9 studies) [121]. Our review includes all these studies, mostly in the categories of aggregated contact or outbreak investigations, with compatible summary estimates (Figure 1). We categorised one report [81] with other studies in which a defined population was screened. The summary estimate in the third systematic review (16%, 95% CI 10–23%, 41 studies) [119] was similar to that of other systematic reviews, despite inclusion of studies with no information about follow-up. In comparison with other reviews, rather than restricting inclusion, we give a comprehensive overview of studies with adequate follow-up, with assessment of risks of bias and exploration of heterogeneity (S2-S7 Figures). The three versions of this review to date have shown how types of evidence change over time, from single family investigations to large screening studies (S1 Table).

### Interpretation

The findings from systematic reviews, including ours [119-121], do not support the claim that a large majority of SARS-CoV-2 infections are asymptomatic [122]. We estimated that, across all study settings, the proportion of SARS-CoV-2 infections that is asymptomatic throughout the course of infection is 20% (95% CI 17–25%). The wider prediction interval reflects the heterogeneity between studies and indicates that future studies with similar study designs and in similar settings will estimate a proportion of asymptomatic infections from three to 67%. Studies that detect SARS-CoV-2 through screening of defined populations irrespective of infection status at enrolment should be less affected by selection biases. In this group of studies, the estimated proportion of asymptomatic infection was 31% (95% CI 26–37%, prediction interval 24–38%). This estimate suggests that other studies might have had an over-representation of participants diagnosed because of symptoms, but there were also potential selection biases in screening studies that might have overestimated the proportion of asymptomatic infections. Our knowledge to date is based on data collected during the acute phase of an international public health emergency, mostly for other purposes. To estimate the true proportion of asymptomatic SARS-CoV-2 infections, researchers need to design prospective longitudinal studies with clear definitions, methods that minimise selection and measurement biases, and transparent reporting. Serological tests, in combination with virological diagnostic methods, might improve ascertainment of SARS-CoV-2 infection in asymptomatic populations. Prospective documentation of symptom status would be required, and improvements in the performance of serological tests are still needed [123].

Our review adds to information about the relative contributions of asymptomatic and pre-symptomatic infection to overall SARS-CoV-2 transmission. Since all people infected with SARS-CoV-2 are initially asymptomatic, the proportion that will go on to develop symptoms can be derived by subtraction from the estimated proportion with true asymptomatic infections; from our review, we would estimate this fraction to be 80% (95% CI 75–83%). Since SARS-CoV-2 can be transmitted a few days before the onset of symptoms [124], pre-symptomatic transmission likely contributes substantially to overall SARS-CoV-2 epidemics. The analysis of secondary attack rates provides some evidence of lower infectiousness of people with asymptomatic than symptomatic infection (Figure 3) [36,65,66,90,111], but more studies are needed to quantify this association more precisely. If both the proportion and transmissibility of asymptomatic infection are relatively low, people with asymptomatic SARS-CoV-2 infection should account for a smaller proportion of overall transmission than pre-symptomatic individuals. This is consistent with the findings of the only mathematical modelling study in our review that explored this question [19]. Uncertainties in estimates of the true proportion and the relative infectiousness of asymptomatic SARS-Cov-2 infection and other infection parameters contributed to heterogeneous predictions about the proportion of pre-symptomatic transmission [20,33,51,63,78,91].

### Implications and unanswered questions

Integration of evidence from epidemiological, clinical and laboratory studies will help to clarify the relative infectiousness of asymptomatic SARS-CoV-2. Studies using viral culture as well as RNA detection are needed since RT-PCR defined viral loads appear to be broadly similar in asymptomatic and symptomatic people [116,125]. Age might play a role as children appear more likely than adults to have an asymptomatic course of infection (Figure 1) [126]; age was poorly reported in studies included in this review (Table 1).

SARS-CoV-2 transmission from people who are either asymptomatic or pre-symptomatic has implications for prevention. Social distancing measures will need to be sustained at some level because droplet transmission from close contact with people with asymptomatic and pre-symptomatic infection occurs. Easing of restrictions will, however, only be possible with wide access to testing, contact tracing and rapid isolation of infected individuals. Quarantine of close contacts is also essential to prevent onward transmission during asymptomatic or pre-symptomatic periods of those that have become infected. Digital, proximity tracing could supplement classical contact tracing to speed up detection of contacts to interrupt transmission during the pre-symptomatic phase if shown to be effective [19,127]. The findings of this systematic review of publications early in the pandemic suggests that most SARS-CoV-2 infections are not asymptomatic throughout the course of infection. The contribution of pre-symptomatic and asymptomatic infections to overall SARS-CoV-2 transmission means that combination prevention measures, with enhanced hand and respiratory hygiene, testing tracing and isolation strategies and social distancing, will continue to be needed.

## Data Availability

All data are fully available

## Supporting Information

S1 PRISMA Checklist

S1 Text. Search strings

S1 Figure. Flow chart

S2 Figure. Review question 1, forest plot of included studies, by study precision

S3 Figure. Risk of bias in studies included in review question 1 and review question 2

S4 Figure: Review question 2, forest plot of included studies, by study precision

S5 Figure: Sub-group analysis, review question 1, comparing studies of hospitalised patients with all other settings

S6 Figure: Sensitivity analysis, review question 1, omitting studies that were preprints at the time of literature search

S7 Figure. Assessment of credibility of mathematical modelling studies

S1 Table. Types of study included in successive versions of the living systematic review, as of 10 June 2020

S2. Table. Location of studies contributing data to review questions 1 and 2

